# Incidence of Guillain-Barré syndrome following SARS-CoV-2 immunization in Mexico: A nation-wide registry of seven COVID-19 vaccines

**DOI:** 10.1101/2022.04.11.22273754

**Authors:** Miguel García-Grimshaw, Javier Andrés Galnares-Olalde, Omar Yaxmehen Bello-Chavolla, Anaclara Michel-Chávez, Arturo Cadena-Fernández, María Eugenia Briseño-Godínez, Neftali Eduardo Antonio-Villa, Isaac Nuñez, Alonso Gutiérrez-Romero, Laura Hernández-Vanegas, María del Mar Saniger-Alba, Roger Carrillo-Mezo, Santa Elizabeth Ceballos-Liceaga, Guillermo Carbajal-Sandoval, Fernando Daniel Flores-Silva, José Luis Díaz-Ortega, Hugo López-Gatell, Ricardo Cortes-Alcalá, José Rogelio Pérez-Padilla, Erwin Chiquete, Gustavo Reyes-Terán, Antonio Arauz, Sergio Iván Valdés-Ferrer

## Abstract

**Background:** Guillain-Barré syndrome (GBS) as an adverse event following immunization (AEFI) against SARS-CoV-2 has been linked to a few (ChAdOx1 nCov-19 and Ad26.COV2-S), but not all vaccines, including mRNA-based ones. Epidemiological information on GBS among recipients of other SARS-CoV-2-directed vaccines among Latinx/Hispanic recipients is scarce.

**Methods:** We report GBS incidence per million administered doses from a nationwide Mexican retrospective registry of adult (≥18 years) recipients of 81,842,426 doses of seven vaccines against SARS-CoV-2 immunized between December 24, 2020, and October 29, 2021. Cases were collected through a passive epidemiological surveillance system and defined as events occurring within 42 days from immunization. Vaccines were analyzed individually and by vector as either mRNA-based (mRNA-1273 and BNT162b2), adenovirus-vectored (ChAdOx1 nCov-19, rAd26-rAd5, Ad5-nCoV, and Ad26.COV2-S), or inactivated whole-virion-vectored (CoronaVac).

**Findings:** We identified 97 patients (52 [53.6%] males; median age 44 years (interquartile range 33–60), for an overall observed incidence of 1.19/1,000,000 doses (95% confidence interval [CI] 0.97–1.45), higher among Ad26.COV2-S (3.86/1,000,000 doses, 95% CI 1.50–9.93) and BNT162b2 (1.92/1,00,000 doses, 95% CI 1.36–2.71) recipients. The overall interval from vaccination-to-GBS symptoms onset was 10 days (interquartile range 3–17). Preceding diarrhea (≤ 4 weeks) was reported in 21.6%, and four (4.1%) more had mild COVID-19. Only 18 patients were tested for Campylobacter jejuni infection; 16 (88.9%) were positive. Electrophysiological examinations were performed in 76 (78.4%) patients (axonal in 46 [60.5%] and demyelinating in 25 [32.8%]); variants were similar between platforms. On initial evaluation, 91.8% had a GBS disability score ≥ 3. Seventy-five (77.3%) patients received intravenous immunoglobulin, seven (7.2%) plasma exchanges, and 15 (15.5%) were treated conservatively. There were 10 (10.3%) deaths, and 79.1% of survivors were unable to walk independently at discharge.

**Interpretation:** In our population, GBS was an infrequent AEFI, observed in less than 1.2/1,000,000 administered doses of vaccines against SARS-CoV-2. Observed incidences were higher among Ad26.COV2.S and BNT162b2 recipients individually and for mRNA-vectored vaccines as a group.

## Introduction

Vaccines are considered potential triggers for Guillain-Barré syndrome (GBS), mainly after the 1976 GBS outbreak among recipients of seasonal influenza A vaccines; however, more recent vaccines are associated with, at most, a small increase in GBS incidence.^1^ GBS is the most frequent cause of acute flaccid weakness, with an incidence of 1.1–1.8 cases per 100,000 person-years worldwide.^2,3^ The epidemiology of adverse events following immunization (AEFI) occurring after SARS-CoV-2 vaccination, including GBS, remains incompletely understood, particularly in underdeveloped countries and underserved regions data on neurologic AEFI comes from a few countries and involves only a handful of vaccines.^4–6^

According to the Mexican General Board of Epidemiology, in 2019 (i.e., pre-COVID-19), Mexico reported a GBS incidence of 0.71 cases per 100,000 person-years.^7,8^ We previously reported a preliminary incidence of GBS ranging from 0.18–0.43 cases per 100,000 doses administered among 3.9 million first-dose recipients of BNT162b2—the only vaccine in use at the time of those reports—which fell within the expected (pre-COVID-19 and pre-SARS-CoV-2 vaccine) incidence.^9,10^ However, as nationwide immunization efforts incorporated more vaccines, epidemiological data from the United States and the United Kingdom suggested epidemiological associations between two adenovirus-vectored vaccines (Ad26.COV2.S [1 case per 100,000 doses administered], and ChAdOx1 nCov-19 (0.87 per 100,000 first-doses administered]) and GBS.^11–13^

At the time of writing this manuscript, there is no data on this potential AEFI among the Latinx population, a heterogeneous group that is commonly underrepresented in clinical trials. Also, in this report, we evaluate GBS following the administration of vaccines used in low-income and middle-income countries.^6,14^ Mexico started its anti-SARS-CoV-2 vaccination program on December 24, 2020. Between December 2020 and September 2021, the Mexican Ministry of Health granted emergency approval for the use of seven different vaccines against SARS-COV-2, using three different platforms: mRNA (mRNA-1273 and BNT162b2), adenovirus (ChAdOx1 nCov-19, rAd26-rAd5, Ad5-nCoV, and Ad26.COV2-S), and inactivated whole-virion (CoronaVac);^15^ thus, being in a unique position to evaluate the differences between multiple of the currently available anti-SARS-CoV-2 vaccines, and not only the commonly used in developed nations for which ample safety information has already been reported.

Here, we report GBS incidence occurring within 42 days after receiving any vaccine against SARS-CoV-2 from a nationwide registry of neurologic AEFI. Also, we report the presence of concomitant well-known GBS potential triggers, clinical presentation, and functional outcomes among recipients of seven different vaccines who sought hospital attention during a 10-month period in Mexico.

## Methods

### Study design and population

Retrospective study of a nationwide registry of Guillain-Barre syndrome (GBS) among recipients of 81,842,426 doses of seven anti-severe acute respiratory coronavirus 2 (SARS-CoV-2) vaccines in Mexico between December 24, 2020, and October 29, 2021. We included hospitalized patients fulfilling the National Institute of Neurological and Communicative Disorders and Stroke clinical features for GBS (Asbury criteria)^16^ who presented during the first 42 days after receiving the last immunization and were officially reported to the Mexican Ministry of Health through a passive epidemiological surveillance system. Patients with missing clinical data, and those with alternative diagnoses explaining the neurological deficits were excluded.

We identified cases using the Mexican epidemiological surveillance system, which collects and processes data on all reported adverse events following immunization (AEFI) from ∼23,300 public and private medical units distributed across the country.^9^ Event severity was initially classified at the local level by the attending medical teams according to the World Health Organization operational case definition as either non-serious (e.g., injection-site pain, swelling, rash, headache, fever, malaise, muscle and/or joint pain) or serious (e.g., put life in danger, require hospitalization, causes disability or death).^17^

Aiming to establish causality, an ad-hoc committee appointed by the Mexican Ministry of Health consisting of five experienced neurologists and a neuroradiologist (A.A, S.I.V.-F, L.E.H.-V., M.M.S.-A., A.G.-R, and R.C.-M.) performed a detailed case-by-case analysis of all potentially serious neurologic AEFI against SARS-CoV-2 through single or multiple virtual sessions with the attending physicians of each patient until causality could be confirmed or ruled out. Operational details of the Mexican epidemiological surveillance system, AEFI definitions, ad-hoc committee case evaluation, and data collection protocols have been previously reported.^18,19^

### Standard protocol approvals, registrations, and patient consents

The study was reviewed and approved by the *Instituto Nacional de Ciencias Médicas y Nutrición Salvador Zubirán* (ID: NER-3903-21-23-1) Ethics and Research Committees who waived the need for signed informed consent due to its observational nature and usage of an anonymized database. This report was elaborated according to the Strengthening the Reporting of Observational Studies in Epidemiology (STROBE) checklist.^20^

### Assessment of potential triggers clinical and electrophysiological features of GBS

Clinical diagnosis was made according to the Asbury criteria.^16^ Clinical variants were determined by the local medical teams and the ad-hoc committee. Disease severity upon admission and at hospital discharge were determined using the GBS disability scale.^21^ Severe disease was defined as a GBS disability scale ≥ 3.^2,22^ Detection and testing for potential triggers such as respiratory tract infections, preceding diarrhoea, detection of *Campylobacter jejuni* by stool real-time reverse transcription–polymerase chain reaction (RT-PCR), or other well-known triggers relied on local medical teams. Due to limited access, testing for anti-ganglioside antibodies was not routinely performed.

The probability of walking independently at six months was estimated using the modified Erasmus GBS outcome score (mEGOS) on the seventh day after admission.^23^ The risk of developing respiratory failure during the first week of admission was evaluated using the Erasmus GBS respiratory insufficiency score (EGRIS).^24^ When available, electrophysiological subtypes were determined locally and confirmed retrospectively by an experienced neurophysiologist using the raw data from the first nerve conduction studies (NCS) according to the Hadden criteria.^25^ Diagnostic certainty was graded according to Brighton Collaboration GBS Working Group criteria.^22,26^

### Data collection

De-identified data were collected into a secure online database using a standardized case report format filled and reviewed by at least two members of the ad-hoc committee during virtual sessions; by consensus, two researchers adjudicated any differences between the primary reviewers (M.G.-G and S.I.V.-F). Data collection included demographics (age and sex); potential triggers including preceding infections; history of or concurrent confirmed SARS-CoV-2 infection by either RT-PCR or antigen testing; type of administered vaccine and, in the case of two-dose schemes, the number of doses received; the interval in days between last vaccine administration and GBS symptoms onset; GBS clinical severity on admission, as well as NCS, and cerebrospinal fluid (CSF) analysis results; immunomodulatory treatments (plasma exchange [PLEX] or intravenous immunoglobulin [IVIg]); requirement of invasive mechanical ventilation (IMV); length of hospital stay; and functional outcome at discharge. The total number of doses administered and reported AEFIs nationwide were obtained from the Mexican Ministry of Health.

### Statistical analysis

For the purpose of analysing the differences between platforms, we evaluated vaccines according to the used vector as mRNA-based (mRNA-1273 and BNT162b2); adenovirus-vectored (ChAdOx1 nCov-19, rAd26-rAd5, Ad5-nCoV, and Ad26.COV2-S); or inactivated whole-virion-vectored (CoronaVac). Age was stratified according to the mEGOS cut-off values. A statistical power calculation was not required since this is a registry-based analysis. Categorical variables are presented as frequencies with proportions, as for continuous variables, after testing for normality with the Shapiro-Wilk test, are reported as median with interquartile range (IQR) or as mean with standard deviation (SD), as appropriate. Some percentages may not add up to 100% due to rounding. Unadjusted observed incidences for each vaccine subtype and platform per 1,000,000 administered doses with 95% confidence intervals (CI) were calculated using the Wilson method.^27^ To evaluate differences in incidence between vaccines subtypes and platforms, we calculated incidence ratios with 95% CI using the lowest observed incidence for each vaccine and platform as the reference value.^28,29^ Analyses were performed using IBM SPSS Statistics version 26 (IBM Corp., Armonk, NY, USA) and figures were created using GraphPad Prism, version 9 (GraphPad Software, La Jolla, CA, USA).

### Data availability

The manuscript provides all the collected data. After approval by the Instituto Nacional de Ciencias Médicas y Nutrición Salvador Zubirán Ethics and Research Committees, de-identified data to replicate our results will be available to qualified researchers upon written request to the corresponding author.

## Results

During the study period, the Mexican Epidemiological Surveillance System processed 31,095 AEFI reports, of which 30,279 (98%) were categorized as non-serious and 816 (2%) as serious. Among the latter, we identified 111 patients with potential GBS; after ad-hoc committee evaluation, an alternative diagnosis was detected in 11 patients (five with functional neurological disorders, three with compressive radiculopathy, two with acute transverse myelitis, and one with an acute ischemic stroke), and were excluded from this report. Due to missing data to establish a clinical diagnosis of GBS, three more were excluded from the analysis altogether (Figure 1).

**Figure 1.**
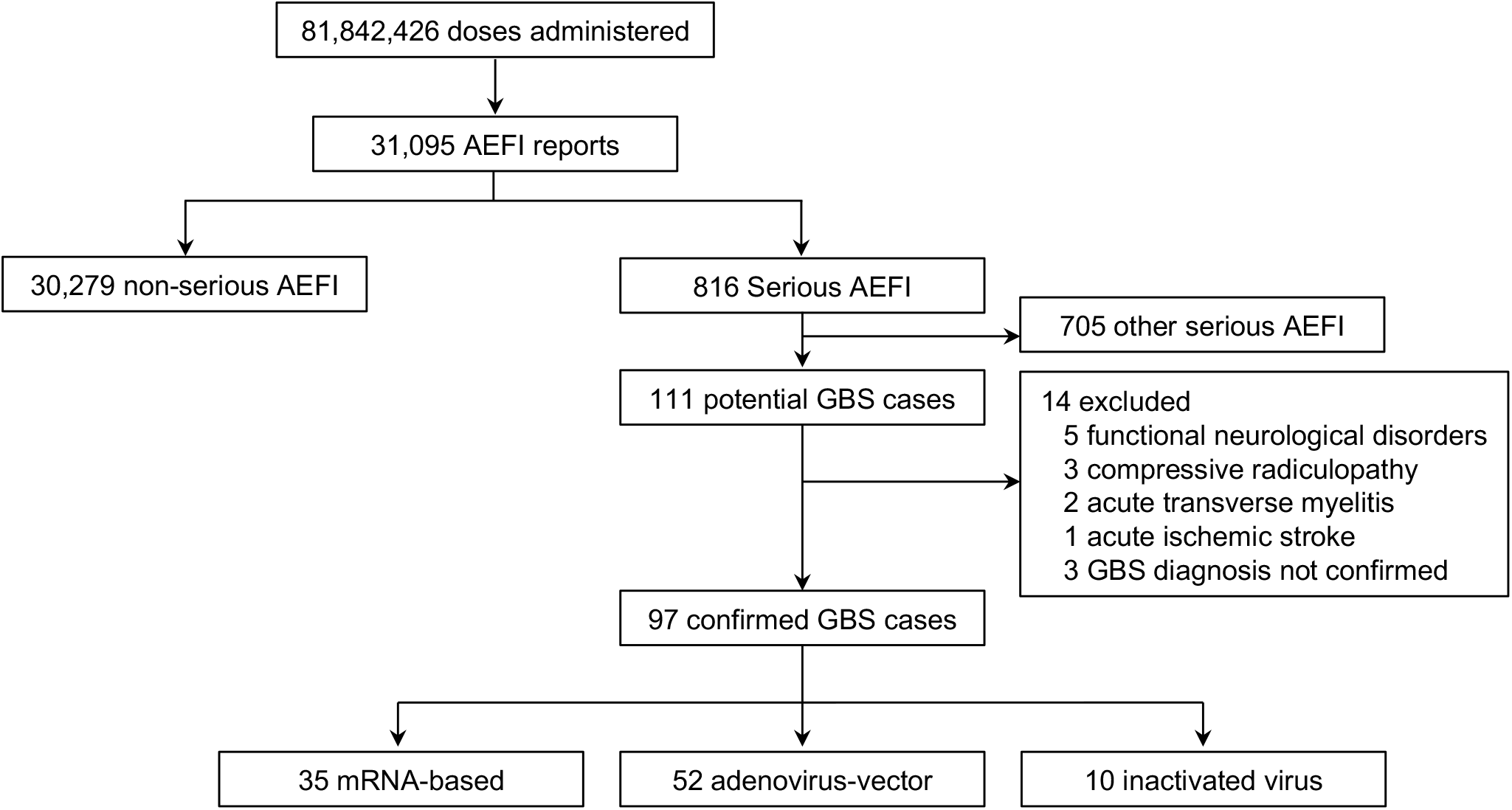
Patient selection flowchart. **Abbreviations:** AEFI, adverse event following immunization; GBS, Guillain-Barré syndrome.

For the final analysis, 97 patients with confirmed GBS were included, representing 11.9% of all serious AEFI. Fifty-two (53.6%) were male; the median age was 44 (IQR 33–60) years (Table 1). Most cases occurred among patients aged 18–40 years with similar proportions between platforms; however, inactivated virus vaccine recipients were older than the total of patients, and those immunized with other platforms with a median age of 59 (IQR 30–63) years. GBS symptoms developed after the first dose in 73 (75.3%) patients and during the first 14 days after the most recent dose in 64 (66%). Figure 2 shows the timing from the last administered dose to GBS symptoms onset according to vaccine platform.

**Table 1.** Baseline characteristics, potential triggers, and clinical presentation according to vaccine platform.

|  | All patients<br>(n = 97) | mRNA-based<br>(n = 35) | Adeno-vector<br>(n = 52) | Inactivated<br>virus<br>(n = 10) |
| --- | --- | --- | --- | --- |
| Sex, n (%) |  |  |  |  |
| Male | 52 (53.6) | 21 (60) | 25 (48.1) | 6 (60) |
| Female | 45 (46.4) | 14 (40) | 27 (51.9) | 4 (40) |
| Age, median (IQR), years | 44 (33–60) | 41 (31–63) | 45 (37–57) | 59 (30–63) |
| Age group, n (%) |  |  |  |  |
| 18–40 years | 41 (42.3) | 15 (42.9) | 22 (42.3) | 4 (40) |
| 41–60 years | 32 (33) | 10 (28.6) | 21 (40.4) | 1 (10) |
| > 60 years | 24 (24.7) | 10 (28.6) | 9 (17.3) | 5 (50) |
| Potential triggers, (%) |  |  |  |  |
| Past SARS-CoV-2 infection | 7 (7.2) | 3 (8.6) | 2 (3.8) | 2 (20) |
| Active SARS-CoV-2 infection | 4 (4.1) | 0 (0) | 2 (3.8) | 2 (20) |
| Diarrhoea, ≤ 4 weeks | 21 (21.6) | 6 (17.1) | 11 (21.2) | 4 (40) |
| Campylobacter jejuni RT-PCR testing | 18 (18.6) | 3 (8.6) | 13 (25) | 2 (20) |
| Positive RT-PCR result* | 16/18 (88.9) | 3/3 (100) | 11/13 (84.6) | 2/2 (100) |
| Most recent vaccine dose, (%) |  |  |  |  |
| First | 73 (75.3) | 23 (65.7) | 43 (82.7) | 7 (70) |
| Second | 24 (24.7) | 12 (34.3) | 9 (17.3) | 3 (30) |
| Days from most recent immunization to GBS symptoms, median (IQR) | 10 (3–17) | 10 (3–21) | 11 (4–19) | 3 (1–15) |
| ≤ 14 days, n (%) | 64 (66) | 24 (68.6) | 32 (61.5) | 8 (80) |
| Neurological symptoms, (%) |  |  |  |  |
| Facial nerve involvement | 24 (24.7) | 8 (22.9) | 16 (30.8) | 0 (0) |
| Bulbar cranial nerves involvement | 30 (30.9) | 11 (31.4) | 18 (34.6) | 1 (10) |
| Weakness in legs only | 20 (20.6) | 7 (20) | 11 (21.2) | 2 (20) |
| Weakness in arms and legs | 74 (76.3) | 28 (80) | 38 (73.1) | 8 (80) |
| Sensory deficits | 46 (47.4) | 15 (42.9) | 29 (55.8) | 2 (20) |
| Clinical variant, n (%) |  |  |  |  |
| Pure motor | 48 (49.5) | 20 (57.1) | 22 (42.3) | 6 (60) |
| Pure sensory | 2 (2.1) | 0 (0) | 2 (3.8) | 0 (0) |
| Sensorimotor | 43 (44.3) | 14 (40) | 27 (51.9) | 2 (20) |
| Miller Fisher syndrome | 4 (4.1) | 1 (2.9) | 1 (1.9) | 2 (20) |
| GBS disability score at admission, n (%) |  |  |  |  |
| 0, 1, or 2 | 8 (8.3) | 3 (8.6) | 5 (9.6) | 0 (0) |
| 3 | 18 (18.6) | 5 (14.3) | 11 (21.2) | 2 (20) |
| 4 | 55 (56.7) | 19 (54.3) | 28 (53.8) | 8 (80) |
| 5 | 16 (16.5) | 8 (22.9) | 8 (15.4) | 0 (0) |
| Erasmus GBS respiratory insufficiency score, median (IQR), points | 4 (3–5) | 4 (3–6) | 4 (3–5) | 4 (3–5) |

\*Proportions for patients tested for *Campylobacter jejuni* by stool RT-PCR.

**Figure 2.**
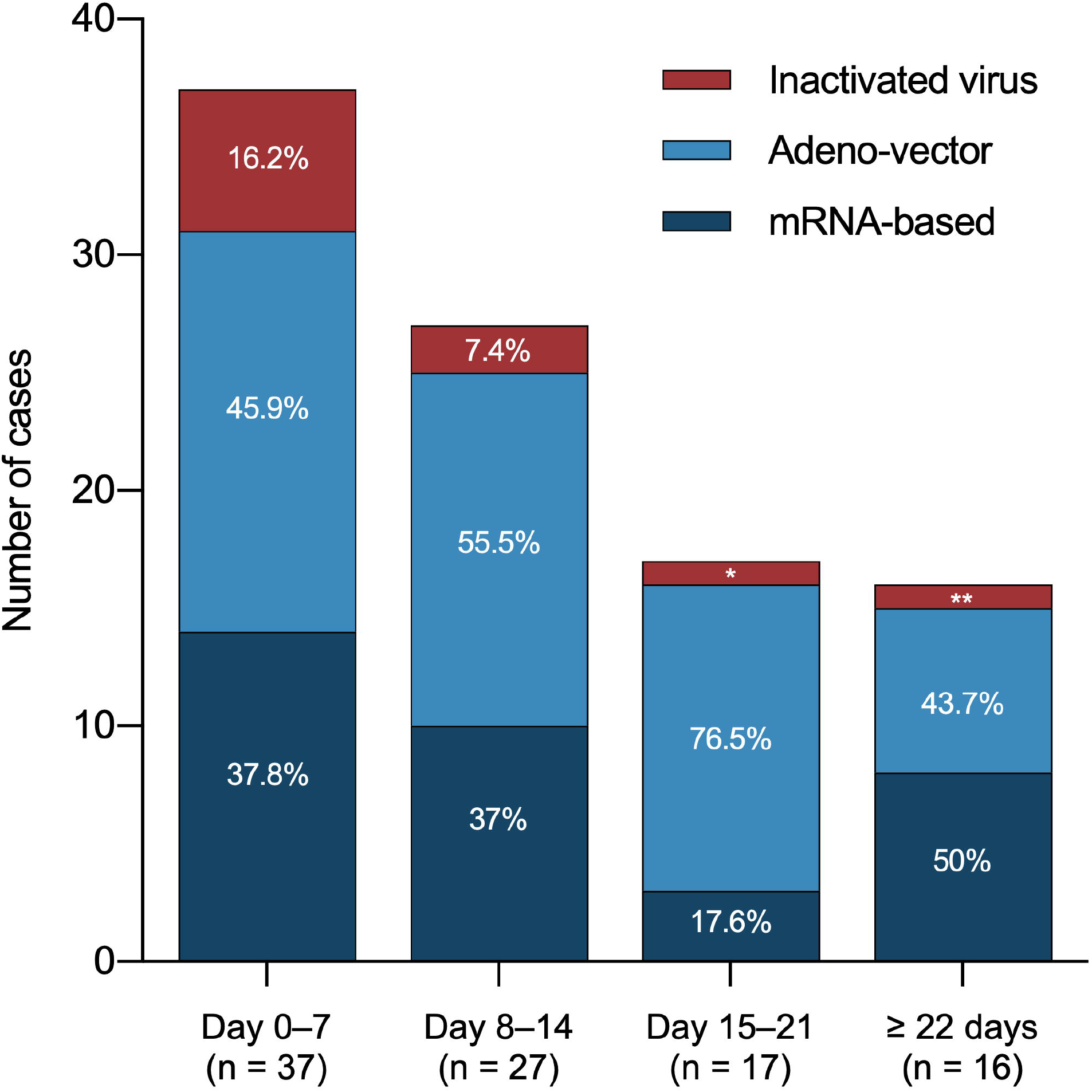
Timing from the last administered dose to Guillain-Barré symptoms onset according to vaccine platform. Inactivated virus includes CoronaVac; adeno-vector includes ChAdOx1 nCov-19, rAd26-rAd5, Ad5-nCoV, and Ad26.COV2-S; mRNA-based includes mRNA-1273 and BNT162b2. *Represents 5.9% of cases occurring during day 15 to 21 after immunization. **Represents 6.3% of cases occurring during ≥ 22 after immunization.

### GBS incidence

The overall observed GBS incidence was 1.19 (95% CI 0.97–1.45) cases per 1,000,000 administered doses (Table 2), with higher observed incidences among recipients of two vaccines: Ad26.COV2-S, 3.86/1,000,000 (95% CI 1.50–9.93) administered doses and BNT162b2, 1.92/1,00,000 (95% CI 1.36–2.71) administered doses. Regarding vaccine platforms, the observed incidence was higher among recipients of mRNA-based vaccines: 1.85/1,000,000 (95% CI 1.33–2.57) administered doses. We then calculated incidence ratios using the CoronaVac (inactivated virus vaccine) as reference value due to its lower incidence. In comparison to CoronaVac, Ad26.COV2-S (5.61/1,00,000; 95% CI 1.76–17.89), BNT162b2 (2.79/1,00,000; 95% CI 1.37–5.68), and the combined mRNA-based vaccines (2.68/1,00,000; 95% CI 1.33–5.42), also had significantly higher incidence ratios (Figure 3 and Supplementary Table 1). Three cases (3.1%) occurred in pregnant women; two occurred during the first trimester (one of them an anembryonic pregnancy) and one during the second trimester, all among first-dose recipients immunized–one each–with BNT162b2, ChAdOx1 nCov-19, or Ad5-nCoV.

**Table 2.** Observed incidence according to vaccine subtype and platform.

| Vaccine | Total doses | Number of cases | Unadjusted incidence (95% CI)* | Vector | Total doses | Number of cases | Unadjusted incidence (95% CI)* |
| --- | --- | --- | --- | --- | --- | --- | --- |
| BNT162b2 | 16,646,623 | 32 | 1.92 (1.36–2.71) | mRNA-based | 18,964,680 | 35 | 1.85 (1.33–2.57) |
| mRNA-1273 | 2,318,057 | 3 | 1.29 (0.44–3.81) |  |  |  |  |
| ChAdOx1 nCov-19 | 38,516,372 | 37 | 0.96 (0.70–1.32) | Adeno-vectored | 48,344,792 | 52 | 1.08 (0.82–1.41) |
| Ad5-nCoV | 2,979,697 | 5 | 1.68 (0.72–3.93) |  |  |  |  |
| rAd26-rAd5 | 5,812,864 | 6 | 1.03 (0.47–2.25) |  |  |  |  |
| Ad26.COV2-S | 1,035,859 | 4 | 3.86 (1.50–9.93) |  |  |  |  |
| CoronaVac | 14,532,954 | 10 | 0.69 (0.37–1.27) | Inactivated virus | 14,532,954 | 10 | 0.69 (0.37–1.27) |
|  |  |  |  | All vaccines | 81,842,426 | 97 | 1.19 (0.97–1.45) |

**Abbreviations:** CI confidence interval. \*Incidence per 1,000,000 doses administered.

**Figure 3.**
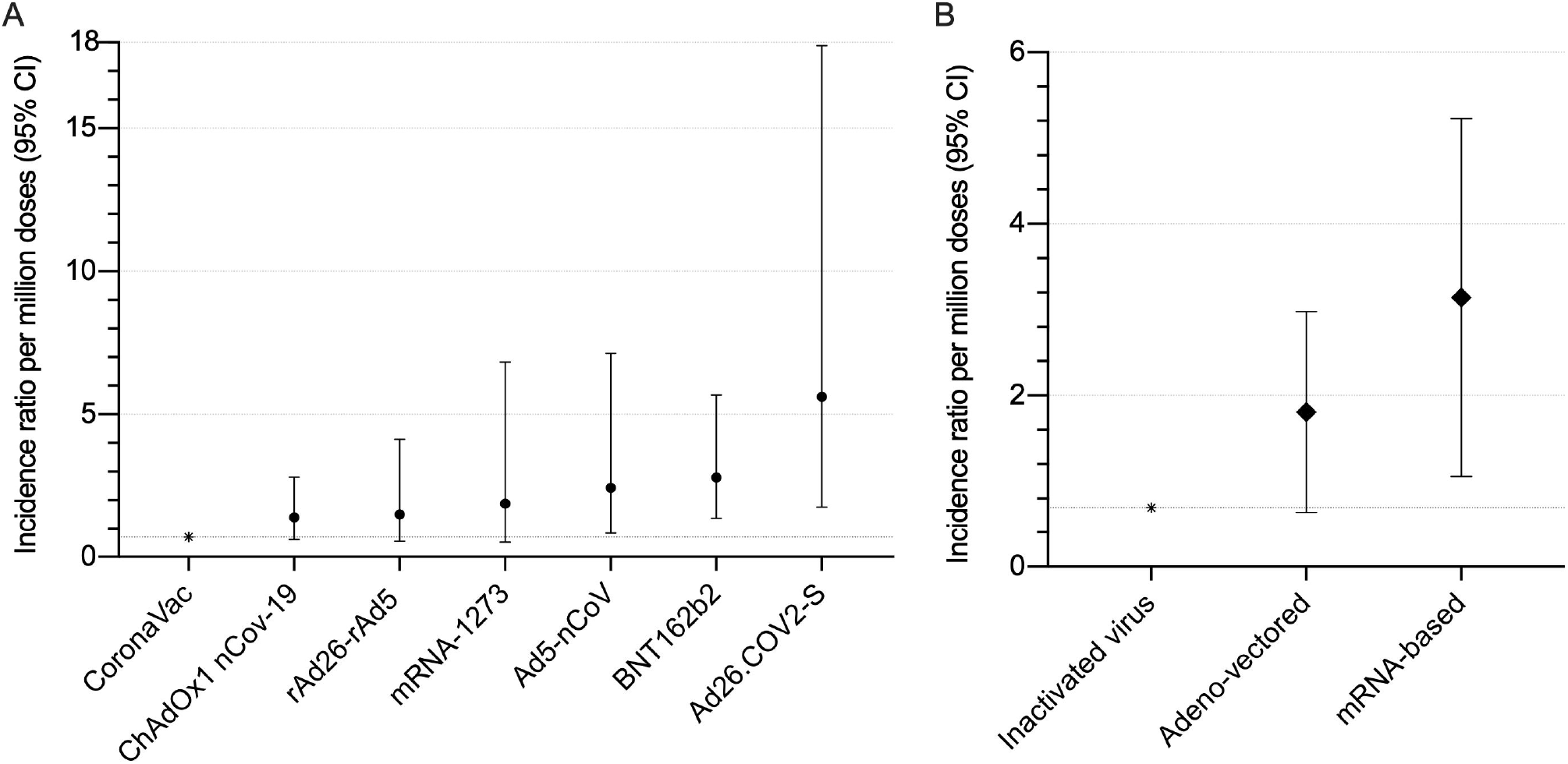
Incidence ratio of Guillain-Barré syndrome according to vaccine subtype and platform. **A**. Incidence ratio according to vaccine subtype. **B**. Incidence ratio according to vaccine platform. All calculations were made using CoronaVac, an inactivated virus single-dose regimen vaccine as the reference. *Reference vaccine and platform value.

### Potential triggers

Twenty-one (21.6%) patients had preceding (≤ 4 weeks) diarrhoea (norovirus was detected in one of them); four had active SARS-CoV-2 infection; of them, three tested positive at the time of GBS symptoms onset, and one four days after; and seven more had a history of COVID-19 (Table 1). In those seven patients, the timing from COVID-19 to GBS symptoms could not be accurately determined. Only 18 patients were tested for *Campylobacter jejuni* infection by stool real-time reverse transcription–polymerase chain reaction (RT-PCR), of whom 16 (88.9%) tested positive. Two patients immunized with the first dose of BNT162b2 had sub-acute (< 30 days) hepatitis A infection. One patient had received influenza immunization 40 days before GBS onset, developing the symptoms three days after receiving the first dose of BNT162b2. None had reports of recent respiratory tract infections. There were no differences in the aforementioned proportions between platforms.

## Clinical and electrophysiologic features

The most frequent presenting signs/symptoms were limb weakness in 74 (76.3%) patients, sensory deficits in 46 (47.4%), cranial (excluding facial) nerve involvement in 30 (30.9%), and facial palsy in 24 (24.7%). On admission, 89 (91.8%) of patients had severe GBS. The most common clinical variants observed were pure motor (49.5%) and sensorimotor (44.3%) (Table 1). In four (4.1%) patients, Miller Fisher syndrome was diagnosed: two after inactivated virus vaccines and one case each after mRNA-based or adenovirus-vectored.

Electrophysiological examinations were performed in 76 (78.4%) patients (Table 3). Among these, 46 (60.5%) had an axonal pattern, 32 (42.1%) had acute motor axonal neuropathy (AMAN), and 14 (18.4%) had acute motor and sensory axonal neuropathy (AMSAN). Twenty-five (32.8%) patients were classified as acute inflammatory demyelinating polyradiculoneuropathy (AIDP) and five (6.6%) as equivocal; none was classified as inexcitable. Cerebrospinal fluid (CSF) analysis was performed in 65 (67%) patients, albuminocytological dissociation was detected in 57 (87.7%). Clinical and electrophysiologic features were similar between vaccine platforms. Fifty (51.5%) patients fulfilled the Brighton level 1 of certainty, 34 (35.1%) level 2 and, 13 (13.4%) level 3. Electrophysiological variants and diagnostic certainty between groups were similar.

**Table 3.**
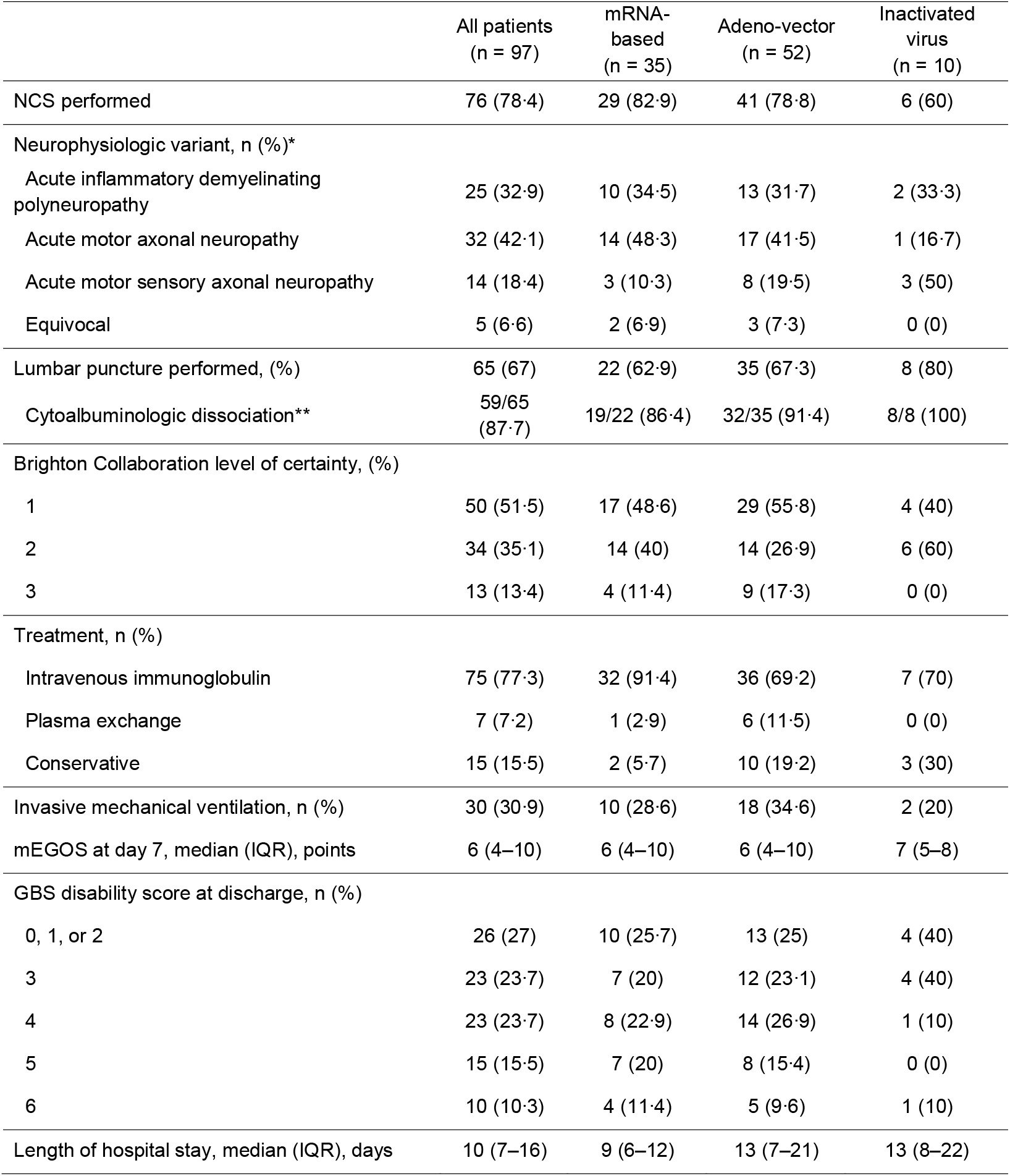
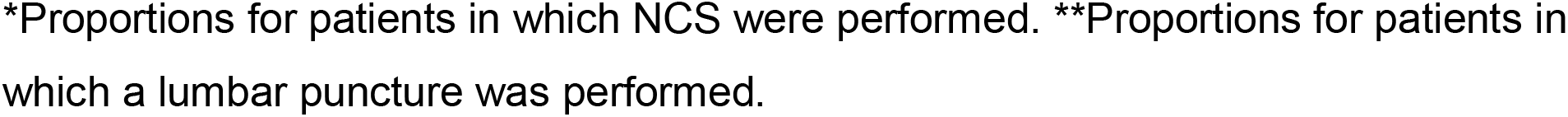
Diagnostic assessment, treatments, and outcomes according to vaccine platform.

|  | All patients<br>(n = 97) | mRNA-based<br>(n = 35) | Adeno-vector<br>(n = 52) | Inactivated<br>virus<br>(n = 10) |
| --- | --- | --- | --- | --- |
| NCS performed | 76 (78.4) | 29 (82.9) | 41 (78.8) | 6 (60) |
| Neurophysiologic variant, n (%)<sup>*</sup> |  |  |  |  |
| Acute inflammatory demyelinating polyneuropathy | 25 (32.9) | 10 (34.5) | 13 (31.7) | 2 (33.3) |
| Acute motor axonal neuropathy | 32 (42.1) | 14 (48.3) | 17 (41.5) | 1 (16.7) |
| Acute motor sensory axonal neuropathy | 14 (18.4) | 3 (10.3) | 8 (19.5) | 3 (50) |
| Equivocal | 5 (6.6) | 2 (6.9) | 3 (7.3) | 0 (0) |
| Lumbar puncture performed, (%) | 65 (67) | 22 (62.9) | 35 (67.3) | 8 (80) |
| Cytoalbuminologic dissociation<sup>**</sup> | 59/65<br>(87.7) | 19/22 (86.4) | 32/35 (91.4) | 8/8 (100) |
| Brighton Collaboration level of certainty, (%) |  |  |  |  |
| 1 | 50 (51.5) | 17 (48.6) | 29 (55.8) | 4 (40) |
| 2 | 34 (35.1) | 14 (40) | 14 (26.9) | 6 (60) |
| 3 | 13 (13.4) | 4 (11.4) | 9 (17.3) | 0 (0) |
| Treatment, n (%) |  |  |  |  |
| Intravenous immunoglobulin | 75 (77.3) | 32 (91.4) | 36 (69.2) | 7 (70) |
| Plasma exchange | 7 (7.2) | 1 (2.9) | 6 (11.5) | 0 (0) |
| Conservative | 15 (15.5) | 2 (5.7) | 10 (19.2) | 3 (30) |
| Invasive mechanical ventilation, n (%) | 30 (30.9) | 10 (28.6) | 18 (34.6) | 2 (20) |
| mEGOS at day 7, median (IQR), points | 6 (4–10) | 6 (4–10) | 6 (4–10) | 7 (5–8) |
| GBS disability score at discharge, n (%) |  |  |  |  |
| 0, 1, or 2 | 26 (27) | 10 (25.7) | 13 (25) | 4 (40) |
| 3 | 23 (23.7) | 7 (20) | 12 (23.1) | 4 (40) |
| 4 | 23 (23.7) | 8 (22.9) | 14 (26.9) | 1 (10) |
| 5 | 15 (15.5) | 7 (20) | 8 (15.4) | 0 (0) |
| 6 | 10 (10.3) | 4 (11.4) | 5 (9.6) | 1 (10) |
| Length of hospital stay, median (IQR), days | 10 (7–16) | 9 (6–12) | 13 (7–21) | 13 (8–22) |

\*Proportions for patients in which NCS were performed. \*\*Proportions for patients in which a lumbar puncture was performed.

### Treatment and outcomes

Treatment and outcomes between vaccine platforms were similar. Eighty-two (84.5%) patients received immunomodulatory treatment: 75 (77.3%) intravenous immunoglobulin (IVIg), and seven (7.2%) plasma exchange (PLEX); 15 (15.5%) patients were treated conservatively (Table 3). None received concomitant steroids, including those four (4.1%) patients with concomitant mild SARS-CoV-2 infection. Thirty (30.9%) required invasive mechanical ventilation (IMV); the median length of stay was 10 days (IQR 7–16). At discharge, 79.1% (61/87) of patients were unable to walk independently (GBS disability score ≥ 3). There were 10 (10.3%) deaths: the cause was septic shock in six patients, dysautonomia in three, and in one (a pregnant woman) due to respiratory failure due to ventilator-associated pneumonia. There were no reports of pulmonary embolism-related deaths.

## Discussion

This analysis of passive epidemiological surveillance monitoring of more than 81.8 million doses of seven anti-SARS-CoV-2 vaccines in Mexico suggests that GBS is an exceedingly rare AEFI, regardless of the vaccine used. Real-world, population-wide analysis is crucial to identify AEFI that may not have been detected in randomized clinical trials. While GBS incidence has been reported for some vaccines, our study is the first one to report the frequency and characteristics of patients who developed GBS as a potential AEFI against multiple SARS-CoV-2 vaccines approved for emergency use in a single Country, allowing us to evaluate the safety of individual vaccines, as well as by vaccine platform.

Since the 1976 swine influenza vaccination campaign in the United States, which prompted the first formal GBS diagnostic criteria.^16,30^ Until the appearance of multiple SARS-CoV-2 vaccines, no clear risk associations had been observed between vaccines and GBS.^1,31–33^ Interestingly, 3.1% of our cases occurred during pregnancy, GBS during pregnancy is considered a rare event, occurring at a rate of 2.8 (95% CI 0.5–9.3) cases per million person-years, and little is known about pregnancy-related immunologic triggers.^34^ Given the number of anti-SARS-CoV-2 vaccines being applied globally with vaccines that were rolled out soon after emergency approval was granted due to the burden of COVID-19, further surveillance for infrequent AEFI is needed.

The overall incidence observed in this study increased 1.67-fold in comparison to the expected (pre-COVID-19) incidence in Mexico.^7,8^ Regarding mRNA-based vaccines, previous reports suggest a lack of association between these vaccines and GBS.^9,10,35–37^ The unadjusted GBS incidence we observed for mRNA-based vaccines is similar to a previous report including recipients of 13,952,901 doses of either mRNA-1273 and BNT162b2.^37^ In the United States, a lower but similar unadjusted incidence between these two vaccines was observed (0.68 and 0.69 cases per 1,000,000 doses, respectively).^38^ Interestingly, we observed that BNT162b2 individually–and mRNA-based vaccines as a group–resulted in a slight increase in GBS risk compared to other vaccines and vectors.

Concerning adeno-vectored vaccines, our results support previous reports suggesting an increased risk among Ad26.COV2.S recipients.^12^ We also observed an increased risk of GBS among Ad26.COV2.S recipients (3.86 per 1,000,000 doses administered); however, this frequency was much lower than that reported in the United States (7.8 per 1,000,000 doses administered).^13^ Among ChAdOx1 nCoV-19 recipients, we observed an incidence of 0.96 cases per 1,000,000 doses administered, which is lower than the incidence reported in the United Kingdom National Immunoglobulin Database (0.87 cases per 100,000 first-doses administered or 8.7 per 1,000,000 first-doses administered).^11^

At the writing of this manuscript we found no reports of GBS associations with the adenovirus-vectored vaccines Ad5-nCoV and rAd26-rAd5, and only one case report of a 76-year-old male diagnosed with GBS two weeks after being immunized with CoronaVac (inactivated virus-vectored).^39^ That may be due to the fact that those vaccines are only being used in a few low-income and middle-income countries, where vaccine numbers are still small and cases potentially under reported.^6,40^ Hence our study provides the first large-scale evidence this AEFI among recipients of Ad5-nCoV, rAd26-rAd5 and CoronaVac.

In line with previous studies, and independently of vaccine type, in our cohort, GBS symptoms started within the first 14 days after immunization and mostly among first-dose recipients.^11,12,37,38^ Regarding disease severity, 91.8% of our patients had a severe form (GBS disability score ≥ 3) compared to the 58.5% of severe cases reported by Keh and colleagues among ChAdOx1 nCoV-19 and mRNA-1273 recipients in the United Kingdom.^11^ This may be due to differences in electrophysiological variants, as patients with axonal variants, known to develop a more severe disease course with worse functional outcomes,^41,42^ accounted for 60.5% of our cases, whereas demyelinating variants accounted for 79.5% of theirs. This can be explained in part by genetic and environmental differences, as demyelinating variants are more frequent in Caucasians, while axonal variants are more frequent in Latin American and Asian populations.^42^ The proportion of axonal variants and mortality rate we observed is consistent with pre-COVID-19 rates, where axonal subtypes accounted for up to 60% of cases with an overall mortality rate as high as 12%.^43–45^

Interestingly, when comparing mRNA-based *versus* adeno-vectored vaccines, we observed a higher incidence ratio for mRNA platforms. These variations in such large samples suggest that genetic and environmental factors may result in increased susceptibility to GBS among recipients of specific SARS-CoV-2 vaccines. However, our data, and that of others, indicates that all seven vaccines evaluated in this report are generally safe concerning the risk of developing GBS.

Mechanism-of-disease is beyond the scope of our manuscript. Hypothetically, immunization-elicited antibodies against SARS-CoV-2 may cross-react with self-antigens expressed in the peripheral nervous system, including Schwann cells and nodes of Ranvier.^46,47^ In the case of mRNA-vectored vaccines, it is also possible that the lipid nanoparticles required to prevent enzymatic degradation of mRNA particles may trigger GBS in genetically- or environmentally-susceptible individuals.^48,49^ Still, a causal relationship between anti-SARS-CoV-2 vaccines and GBS is unknown.

While only 18.6% of our cases were evaluated for *Campylobacter jejuni*, more than 90% of those tested positive, and 20% of all patients had preceding diarrhoea, suggesting that other well-known GBS triggers may be the cause and that these cases were coincident with, but unrelated to, SARS-CoV-2 vaccination. Although a trigger cannot be identified in up to one-third of patients with GBS,^42,50^ a comprehensive approach for known triggers must be performed to establish causality accurately, something that should improve in light of our findings.

This report has strengths and limitations. Among the strengths of our manuscript, we relied on an unusually large population of vaccine recipients and included vaccines for which no safety data related to GBS has been reported, including Ad5-nCoV, rAd26-rAd5, and CoronaVac. We would also like to acknowledge some limitations that must be considered to adequately interpret our results. First, interpretation of the study is limited by its descriptive nature. Second, we were unable to estimate incidence rate ratios or adjust incidences by age, sex, or calculate an incidence during pregnancy because we could not obtain the number of administered doses per month, sex, or age group. Third, as AEFI reports rely upon local health care providers, we could not establish causality or accurately determine other relevant clinical data, such as the development of dysautonomia, due to a lack of standardized diagnostic protocols. Fourth, due to the passive nature of the Mexican epidemiological surveillance system, which is less likely to detect cases than active surveillance systems–due to healthcare seeking bias–and because we only included patients evaluated by the ad-hoc committee, our data is susceptible to selection bias. Finally, in line with the former, mildly symptomatic patients presenting (GBS disability score < 2) presenting with non-disabling symptoms or sequelae may be underdiagnosed or underreported, as well as those occurring in rural settings with limited access to medical services.

In conclusion, here we show that GBS is extremely infrequent among recipients of all vaccines against SARS-CoV-2. However, we observed increases in observed frequency among recipients of Ad26.COV2.S and BNT162b2 individually, and mRNA-vectored vaccines as a group, the magnitude of the increase in risk pale in comparison to the magnitude of protection against severe and lethal forms of COVID-19.

## Data Availability

All data produced in the present work are contained in the manuscript

## Author contributions

M.G.-G., J.A.G.-O., A.A., G.R.-T., and S.I.V.-F. jointly led the study conceptualization, development of the research questions, and study design. M.G.-G., I.N., F.D.F.-S., and E.C. performed the analyses. M.G.-G., J.A.G.-O., A.M.-C., A.C.-F., A.A., and S.I.V.-F. wrote the first draft of the paper. A.G.-R., L.H.-V., M.M.S.-A., R.C.-M., S.E.C.-L., G.C.-S., F.D.F.-S., J.L.D.-O., H.L.-G., R.C.-A., J.R.P.-P. data approvals, undertook the data specification and data linkage and provided critical feedback on the manuscript drafts. O.Y.B.-C., and N.E.A.-V., conducted the replication analyses. O.Y.B.-C., N.E.A.-V., I.N., E.C., F.D.F.-S., J.L.D.-O., H.L.-G., R.C.-A., J.R.P.-P., S.E.C.-L., and A.A. contributed to the discussion on protocol development and provided critical feedback on drafts of the paper. All authors approved the protocol, contributed to the critical revision of the paper, and approved the final version of the paper.

## Supplementary Material

**Supplementary Table 1.**
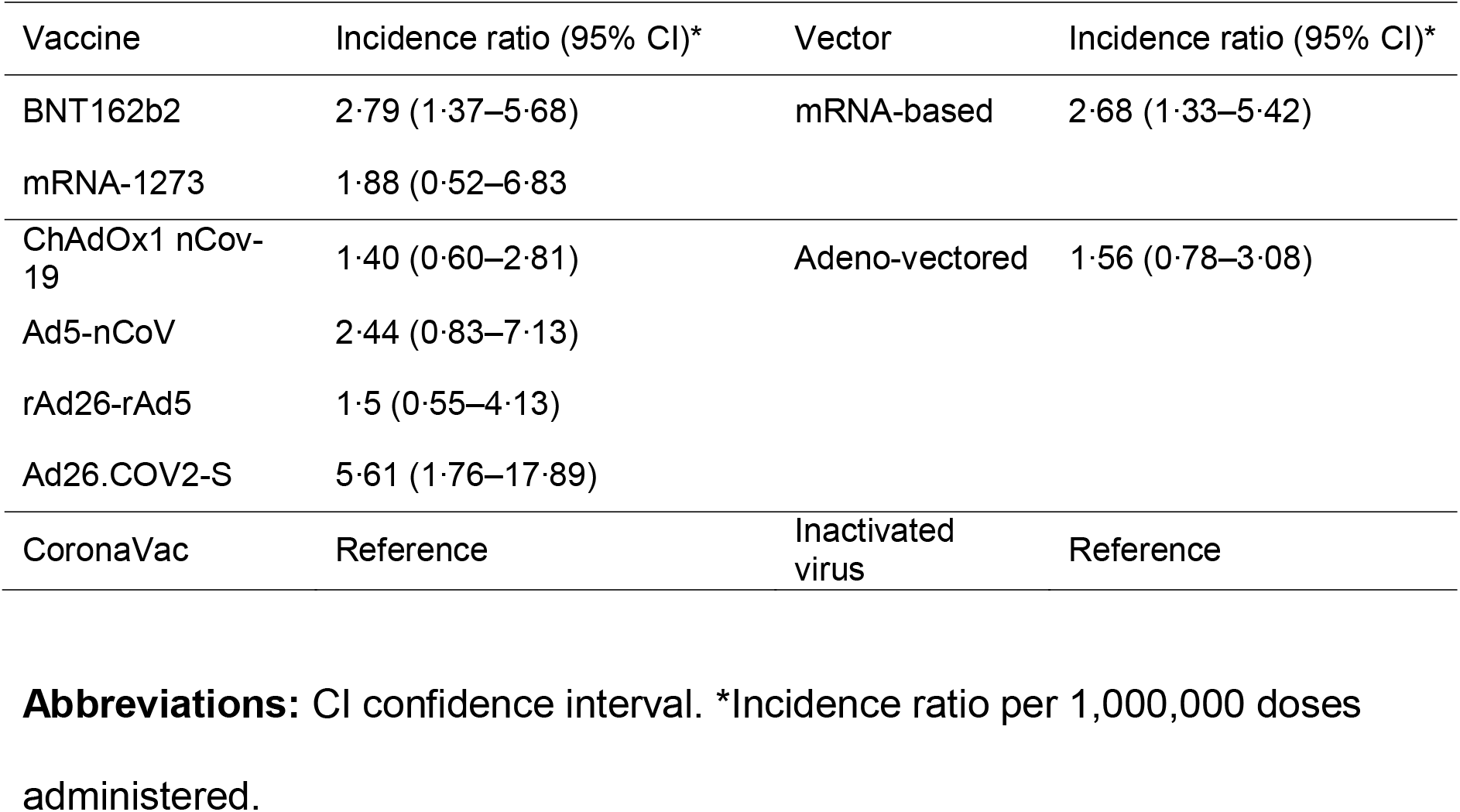
Guillain-Barré Syndrome Incidence Ratio per Vaccine and Vector.

